# Fat infiltration in the vastus medialis implicates joint structural abnormalities in early-stage symptomatic knee osteoarthritis

**DOI:** 10.1101/2022.05.27.22275636

**Authors:** Atsushi Hoki, Ella D’Amico, Fabrisia Ambrosio, Tsubasa Iwasaki, Yoshikazu Matsuda, Hirotaka Iijima

## Abstract

**Objective:** Early knee osteoarthritis (KOA) presents as minor structural abnormalities in joint tissues, such as cartilage and subchondral bone, that cannot be assessed radiographically. Identification of a sensitive and convenient marker for early disease has the potential to enhance patient outcomes. This study determined 1) whether fat infiltration in muscle (i.e., muscle quality), as measured by ultrasound, is associated with structural abnormalities seen in early KOA and 2) which quadriceps muscles are appropriate as a novel marker for early KOA.

**Methods:** Participants with early symptomatic KOA (Kellgren Lawrence grade 1-2) underwent ultrasound assessment to measure the echo intensity of the vastus medialis and rectus femoris. The echo intensity corrected for ultrasound wave attenuation caused by subcutaneous fat was then calculated (i.e., corrected echo intensity). Structural abnormalities were assessed using the whole-organ magnetic resonance score (WORMS). A generalized linear mixed model was used to assess the relationship between the corrected echo intensity and WORMS score.

**Results:** Forty-nine participants (ages: 44-78 years, 65.3% women) with 52 knees were included. After adjustment for covariates, increased corrected echo intensity (i.e., poor muscle quality) in the vastus medialis muscle was significantly associated with greater structural abnormalities, including disrupted cartilage integrity in the medial tibiofemoral joint. The association was not significant in the rectus femoris muscle.

**Conclusion:** Individuals with poor muscle quality in the vastus medialis displayed compromised joint integrity. This study suggests that fat infiltration in vastus medialis assessed by ultrasound is an indicator of early symptomatic KOA.

## INTRODUCTION

Knee osteoarthritis (KOA) is a progressively painful and debilitating condition that can limit activities of daily living(1). Due to a lack of effective disease modifying treatments for KOA, increased clinical interest has been directed towards early detection so as to minimize disease progression(2). While radiography is the current gold standard for assessment of KOA, by the time radiography detects narrowing of the joint space, more than 10% of the cartilage has already been lost, and the subchondral bone area has increased by 15%(3). More recent studies have shown that in patients with early KOA, inflammation of the synovium precedes radiographic signs(4). The identification of sensitive and convenient early-stage markers of whole joint structural abnormalities would expand opportunities for effective disease management.

One promising candidate for identification of early disease is skeletal muscle fatty infiltration (i.e., myosteatosis). It is well established that myosteatosis causes muscle weakness(5), a major risk factor for incident of KOA(6). In addition, myosteatosis elicits endocrine effects via the secretion of adipokines implicated in the pathogenesis of KOA(7). A meta-analysis showed that myosteatosis in the quadriceps muscle is increased in individuals with mild-to-moderate KOA when compared to healthy older adults(8). Further implicating the contribution to disease progression, a longitudinal study reported that myosteatosis in the quadriceps muscle, particularly vastus medialis (VM) muscle, was associated with greater cartilage loss and bone marrow lesions in patients with mild to moderate KOA(9). These previous studies used magnetic resonance imaging (MRI) to assess myosteatosis in quadriceps muscle. However, MRI is both costly and requires long procedure times. Given these barriers, ultrasound represents an attractive alternative to assess myosteatosis in quadriceps. Echo intensity (EI), measured using ultrasound, has previously been established as an accurate and reliable indicator for myosteatosis in comparison to similar measurements using MRI and computed tomography (CT) (10, 11). However, whether increased EI (i.e., poorer muscle quality) of quadriceps muscle can be an indicator of structural abnormalities in earlier stage of KOA has not been assessed.

This cross-sectional study tested the hypothesis that poorer quadriceps muscle quality assessed by ultrasound is associated with greater structural abnormalities of the knee in individuals with earlier stage of symptomatic KOA. Specifically, we posited that EI of the VM muscle is associated with increased structural abnormalities (e.g., cartilage loss, bone marrow lesion), as identified by MRI. This study primarily focused on the VM muscle given that poor muscle quality in people with KOA has predominantly been confirmed only in this muscle(12). Given that some studies have also targeted the rectus femoris muscle (RF) for the muscle quality assessment(8), this study also determined whether structural abnormalities of the knee are associated with poor muscle quality in the RF. Ultimately, we aimed to identify the most appropriate target muscle for the prediction of earlier stage of KOA. The findings of this study support ultrasound-based assessment of VM quality as an indicator of early stage of KOA and represent a promising first step in the development of an effective intervention for KOA.

## METHODS

### Subjects

Individuals with KOA were recruited at the outpatient department of the Matsuda Orthopedic Clinic in Saitama, Japan, from June 2021 to October 2021. All recruited participants were previously seen in the orthopedic clinic for pain in one or both knees and had a diagnosis of KOA within up to 3 months. The presence of KOA was confirmed by an experienced orthopedic surgeon using the radiographically defined Kellgren and Lawrence grading scale (KL grade)(13). Participants also underwent MRI of their symptomatic knee(s). The eligibility criteria were as follows: (1) mild KOA with KL grade 1-2; (2) absence of osteonecrosis of the medial femoral condyle; (3) no history of surgery to the extremities (except intraarticular knee injections); (4) ability to walk without an assistive device; (5) no cognitive dysfunction; (6) no neuromuscular disease; and (7) no trauma to bilateral lower extremities. All these criteria were assessed based on information in the medical records. Given that radiographically defined KL grade 1, but not grade 0, predicts development of KOA to a grade 2 or higher within 3–5 years(14, 15), this study included individuals with KL grades 1 but excluded grade 0. This study was approved by the Ethics Committee of the Medical Corporation Nagomi (No. 20210427). All participants provided signed informed consent.

### Measurements

Muscle quality of the RF and VM was quantified by ultrasound. Structural abnormalities were identified using MRI. These assessments were limited to the symptomatic knee(s), as defined by the presence of knee pain and/or functional limitations from medical examination by physician. Demographic characteristics, body mass index (BMI), ultrasound-based muscle thickness, and self-reported measures of knee pain and disability were also assessed as participant characteristics and/or covariates.

### Ultrasound imaging of RF and VM muscles

Ultrasound images of the symptomatic knee(s) were acquired using a B-mode imaging device that was equipped with an 8-MHz linear-array probe (SNiBLE, Konica Minolta Ltd., Japan). Owing to the lack of evidence for specific ultrasound parameters in the assessment of muscle quality in the RF and VM muscles, ultrasound settings were optimized to visually discriminate the fascia of the RF and VM from the muscle fibers and other surrounding tissues (frequency: 9 MHz, gain: 25 dB, dynamic range: 60), in accordance with a previous study(12). These ultrasound settings were kept consistent across all participants, and images were taken in a relaxed supine position with the arms and legs extended. The RF was evaluated on the transverse line halfway between the anterior superior iliac spine and proximal end of patella while the VM was evaluated at the 5cm medial side from the 20% distal point of anterior superior iliac spine and proximal end of patella(12) (**Figure 1A**). The head of the probe was maintained perpendicular to the muscles examined. A water-soluble transmission gel (PROJELLY, Jex Inc., Japan) was applied to the skin to enhance acoustic coupling. EI of the captured ultrasound images was then assessed using ImageJ software (version 1.53, National Institutes of Health, USA)(16). Regions of interest were selected at depths of 1.0-4.0 cm from the surface of skin on the screen display. The EI of the analysis region was calculated using grayscale values from 0 (black) to 255 (white), with a higher mean pixel intensity value indicating fatty infiltration and/or fibrosis tissue(11, 17) (i.e., poor muscle quality). All ultrasound imaging and ImageJ analyses were performed by a trained investigator (AH). To assess the reliability of the imaging procedure, the same investigator performed another assessment more than six months after the first assessment, revealing an excellent intraclass correlation coefficient (ICC_1,1_: 0.948, 95% confidence interval [CI]: 0.912–0.970)(18). To account for the effect of subcutaneous fat thickness on echo measurements, a correction factor was applied to echo measurements (Equation 1)(11), as recommended in several studies(19, 20). The formula for the corrected EI was as follows:

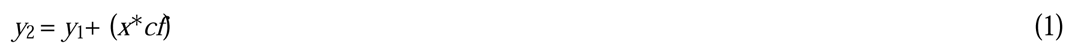

**Figure 1.**
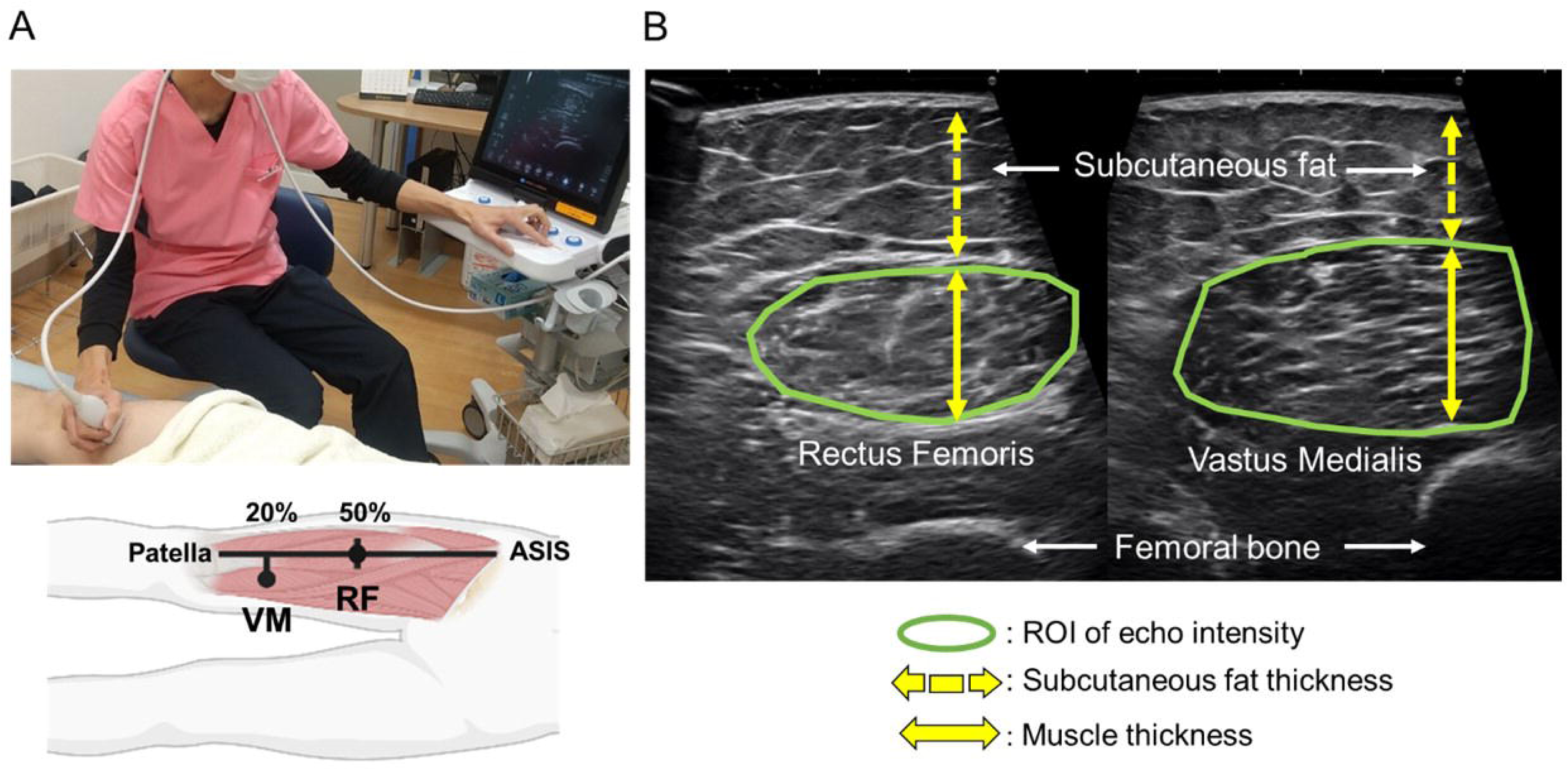
Procedure of ultrasound assessment on quadriceps muscle quality. Example of the data collection procedure in rectus femoris and vastus medialis with ultrasound (**A**). Example of image analysis for the determination of EI, muscle thickness, and subcutaneous fat thickness (**B**). ROIs for EI (solid line circle) were selected at a depth of 1.0-4.0 cm to avoid the surrounding tissue. Muscle thickness (solid line arrow) and subcutaneous fat thickness (dashed line arrow) were measured using an electric caliper. *Abbreviations: ASIS, anterior superior iliac spine; EI, echo intensity; RF, rectus femoris; ROI, region of interest; VM, vastus medialis*.

with *y*_2_= corrected echo intensity; *y*_1_= raw echo intensity; *x*= subcutaneous fat thickness; and *cf*= correction factor of 40.5278.

Subcutaneous fat thickness was measured from the superior border of the superficial aponeurosis to the inferior border of the dermis layer, per the established protocol validated by MRI and computed tomography(11, 21). In addition, as a covariate, muscle thickness (cm) was measured from the cortex of the femur to the superior border of the superficial aponeurosis of the RF and VM muscles(22). The image analysis procedure for EI, subcutaneous fat thickness, and muscle thickness is illustrated in **Figure 1B**.

### Assessment of structural abnormalities in the knee joint by MRI

Structural abnormalities in the knee joint were identified using MRI (1.5 Tesla whole-body MR system, Canon Medical Systems, Japan). Images were obtained as sagittal proton-density-weighted images (3600 msec TR (Repetition Time), 30 msec TE (Echo Time), 256×384 matrix, 3.5 mm slice thickness, 0.3 mm inter-slice gap, 180 mm×180 mm field of view, TSE fact (Turbo Spin Echo fact): 15, scan time: 3 min, frequency encoding: anterior-posterior). We used a semi-quantitative scoring method of the whole-organ magnetic resonance imaging score (WORMS)(23), which has been used previously for individuals with early KOA(24). WORMS includes the following categories: (1) cartilage signal and morphology, (2) subarticular bone marrow abnormality, (3) subarticular cysts, (4) subarticular bone attrition, (5) marginal osteophytes, (6) medial and lateral meniscal integrity, (7) anterior and posterior cruciate ligament integrity, (8) medial and lateral collateral ligament integrity, (9) synovitis, loose bodies, and (10) periarticular cysts/bursae. The whole knee joint was divided into the medial tibiofemoral joint, lateral tibiofemoral joint, and patellofemoral joint. The femoral articular surface, medial tibial plateau and lateral tibial plateau were divided into three sub-regions (anterior, central, posterior). The tibia has an additional subspinous sub-region comprised of the non-articulating portion of the tibial plateau beneath the tibial spines. The assessment was performed by the same investigator (AH) who had undergone three weeks of formal training by an experienced orthopedic surgeon and radiologist prior to the start of the study. The investigator read MRI images in a random order with a blinded subject ID. At the second evaluation of WORMS by the same investigator, more than six months after the initial evaluation, the intraclass correlation coefficient was good (ICC_1,1_: 0.849, 95%CI: 0.752, 0.910).

### Participant characteristics and covariates

Data on participant age, sex, and height were self-reported in the electronic medical records. Body mass was measured on a digital scale with the participants dressed but not wearing shoes. BMI was calculated by dividing body mass (kg) by height (m^2^). A knee radiograph of the symptomatic knee in the anteroposterior view in the weight-bearing position was obtained for all subjects. Using this image, we assessed the femur-tibia angle (FTA) as a measure of the anatomical axis (higher values indicate more varus alignment). The center of the FTA was defined from three points: a point at the base of the tibial spines, at bisecting the femur and tibia, originating 10 cm from the knee joint surface(25). FTA was then corrected to account for sex differences in knee alignment, which was calculated by the addition of 3.5° for women and 6.4° for men, respectively(26). This corrected FTA was used for the subsequent statistical analyses as an alternative measure of full-limb radiograph in weight-bearing position(27).

To quantify knee symptoms, all subjects completed the Knee Injury and Osteoarthritis Outcome Score (KOOS)(28). The KOOS has the following subcategories: pain, symptoms, activities of daily living (ADL), sport and recreation function, and knee-related quality of life, with higher scores (0–100) representing better function. This study used the KOOS pain and ADL subscales for descriptive analysis.

### Statistical analysis

The minimum sample size required to detect significance was calculated from pilot data of individuals with KOA (n = 20) using Power and Sample Size Program, version 3.1.6 (Vanderbilt University Medical Center, USA)(29). Preliminary data indicated that the standard deviations of the WORMS and EI were 9.415 and 17.575, respectively, with a slope estimate of 0.209 obtained when WORMS was regressed against EI. As such, 46 subjects were needed to reject the null hypothesis that this slope equals zero with a probability (power) of 0.8. The Type I error probability associated with this test of null hypothesis was 0.05. The same approach identified that 38 subjects would be needed to detect a significant relationship between the

WORMS and corrected EI. After considering the potential 10% dropout rate due to the exclusion criteria and missing data, the required sample size of this study was 51 participants. To account for the similarity between the right and left knees, a generalized linear mixed model, with WORMS total score as fixed effects and patient ID as random-effect intercept and slope, was used to assess the relationship between the WORMS (continuous variable) and EI of VM (continuous variable) or corrected EI of VM (continuous variable). Age (continuous variable), muscle thickness (continuous variable), and sex were included as covariates. These covariates were chosen based on their potential correlation with joint structural abnormalities and muscle quality(30, 31). BMI was not considered as a covariate because of a possible strong correlation with muscle thickness(32). To address the possibility that structural abnormalities might be associated with changes in tissue quality in muscles other than the VM, we repeated the generalized linear mixed model including EI or corrected EI of RF muscle. Since the WORMS total score includes structural abnormalities of whole knee joint tissues (i.e., cartilage, subchondral bone, ligaments, menisci, and synovium), we performed a sensitivity analysis focusing on the WORMS sub-score of cartilage at medial tibiofemoral joint. In the sensitivity analysis, we repeated the generalized linear mixed model described above.

All statistical analyses were performed using EZR, version 1.54 (Saitama Medical Center, Jichi Medical University, Japan), which is a graphical user interface for R, version 4.0.3 (The R Foundation for Statistical Computing, Austria), a modified version of R commander designed to add statistical functions frequently used in biostatistics. Statistical significance was set at *p* <0.05.

## RESULTS

A total of 51 participants (54 knees) were recruited. Two participants displayed severe KOA with KL grade 3 and were therefore excluded. Therefore, 49 participants (age: 44-78 years, 65.3% women) with a total of 52 symptomatic knees were ultimately included. **Table 1** summarizes the participants’ characteristics. Of the 49 participants with mild KOA, 30 (61%) had a normal BMI (BMI 18.5 to 24.9 kg/m^2^), 14 (29%) were pre-obese (BMI 25.0 to 29.9 kg/m^2^), and 5 (10%) were obese (BMI <30 kg/m^2^). Of the 52 knees, 39 (75%) had varus alignment (corrected FTA >181°), 12 (23%) had neutral alignment (178° < corrected FTA <181°), and 1 (2%) had valgus alignment (corrected FTA <178°)(26).

**Table 1.**
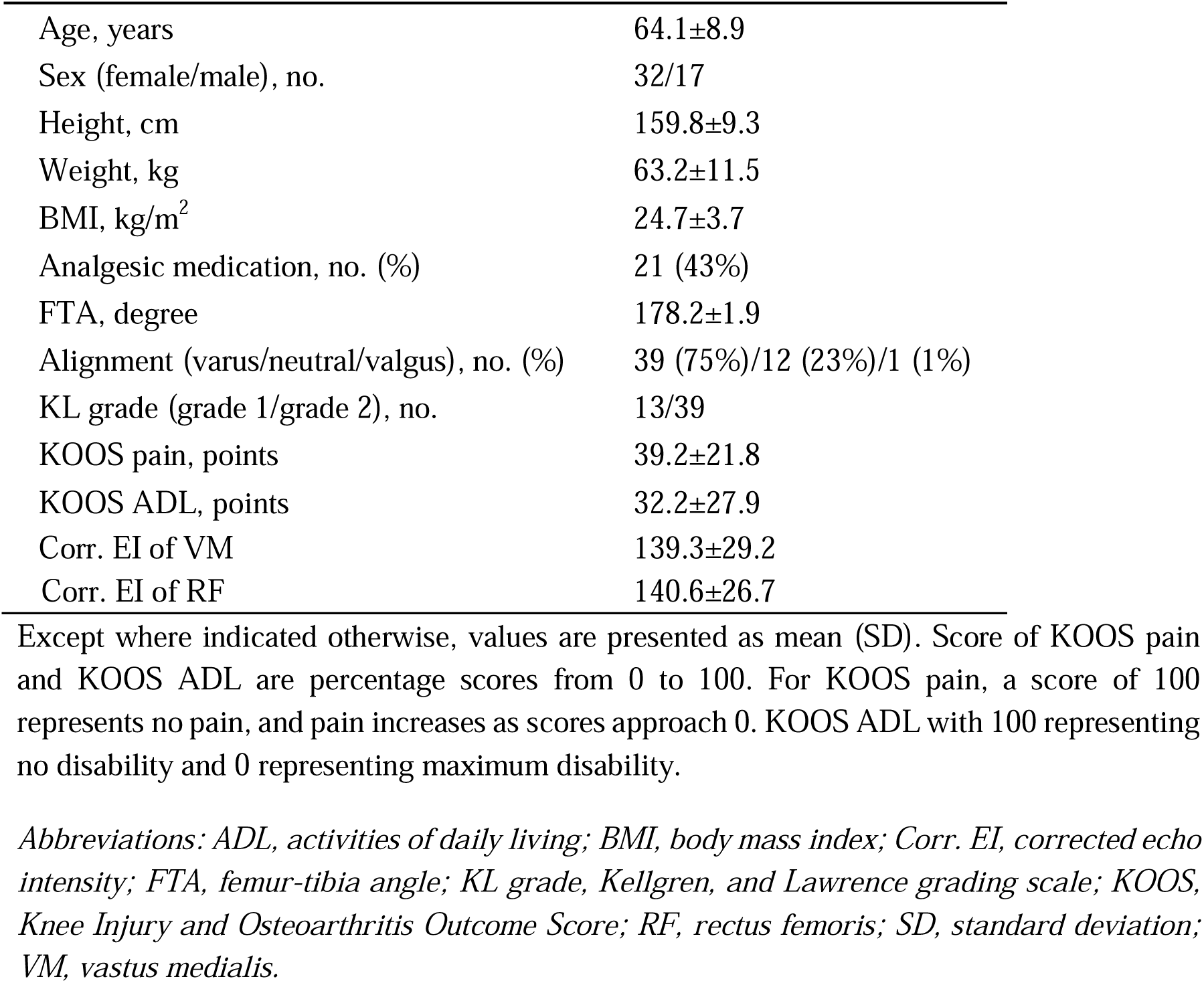
Characteristics of the study subjects (49 participants with 52 symptomatic knees)

### Individuals with poorer VM muscle quality display higher disruption in cartilage integrity

Consistent with the findings seen with early-stage symptomatic KOA(24), 52 knees (100%) had cartilage defects, 51 (98%) had meniscal tears, and 21 (40%) had bone marrow lesions in the WORMS subcategories. Among these subcategories, the cartilage sub-score in the medial compartment of the tibiofemoral joint showed the highest median score (**Figure 2**), where was most likely to have relations to KOA(33). Ultrasound and MRI assessments revealed inter-subject variability in quadriceps muscle quality, as identified by ultrasound assessment, as well as structural abnormalities of the cartilage, as identified on MRI assessment among individuals with mild KOA (**Figure 3**). Since all included participants displayed mild KOA with KL grade 1-2, the identification of inter-subject variability in structural abnormalities highlights the ability of MRI to assess structural abnormalities in the earlier stages of KOA. The generalized linear mixed model revealed that the poor muscle quality of the VM, as assessed by corrected EI, was significantly associated with higher joint structural abnormalities after adjusting for muscle thickness, age, and sex. Decreased muscle quality of the RF was also associated with increased joint abnormalities, though the trend was blunted as compared to VM (**Table 2**). The corrected EI of the VM had higher predictive ability for joint structural abnormalities (r^2^=0.44, p=0.003) compared to uncorrected EI of VM (r^2^=0.39, p=0.02) (**Figure 4A-B**).

**Figure 2.**
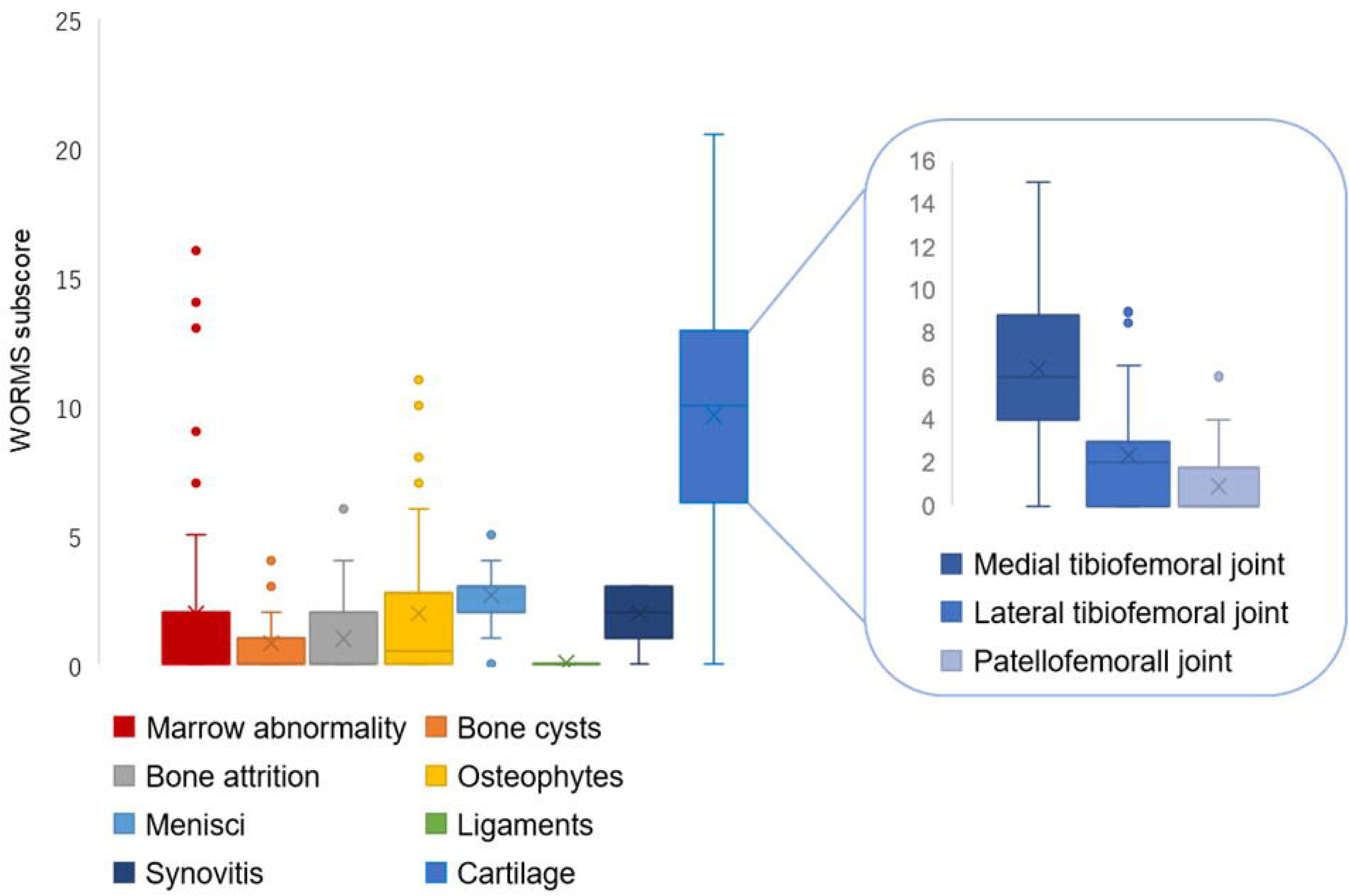
The distribution of WORMS sub-score. The vertical axis shows sub-score in each subcategory. The box plot shows the interquartile range. The inset graph right above shows the breakdown of cartilage sub-score at each joint in the knee. *Abbreviations: WORMS, whole-organ magnetic resonance imaging score*.

**Figure 3.**
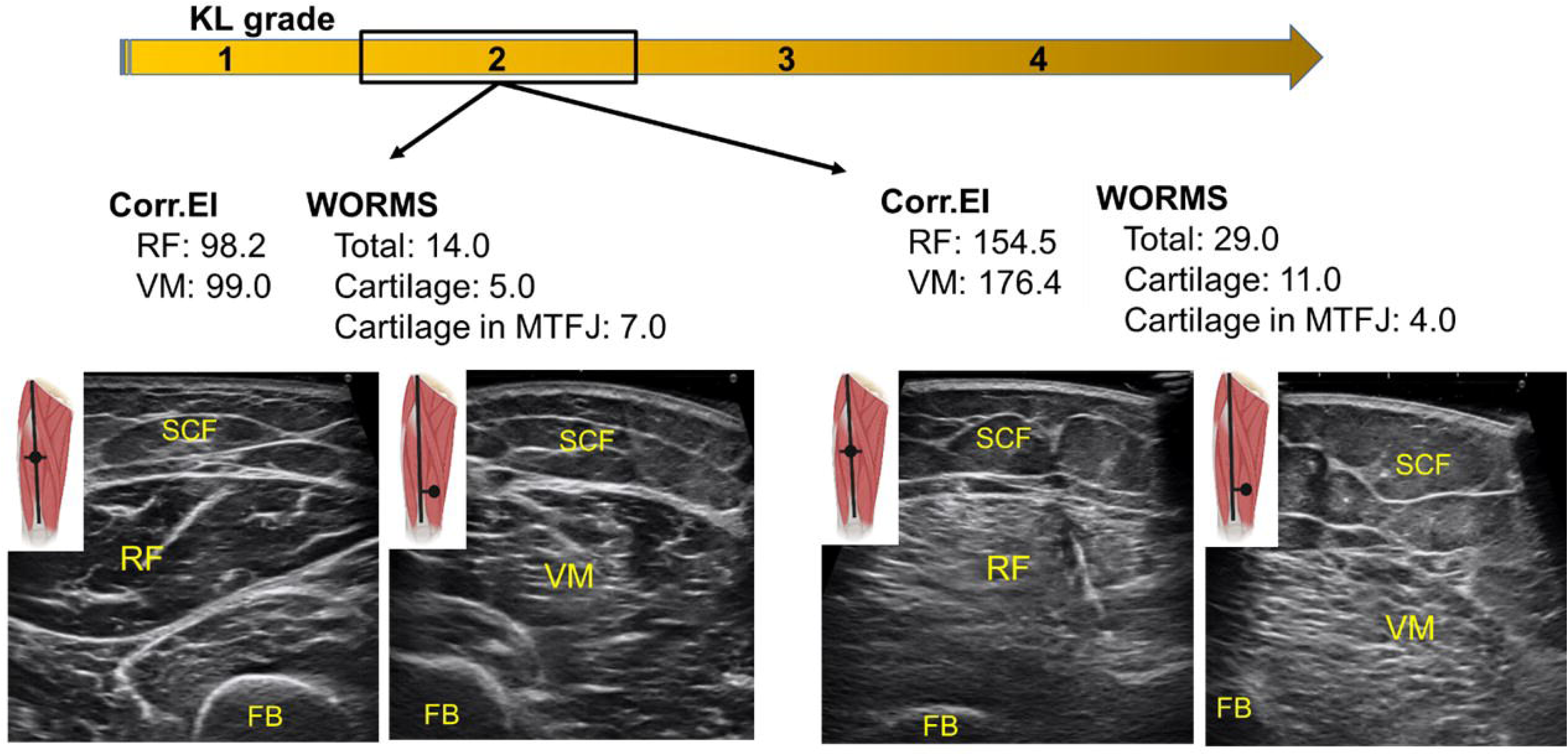
Representative ultrasound images of quadriceps muscle. Images show low corrected EI and low WORMS in her early 60’s woman with KL2 (two images on the left) and high corrected EI and high WORMS in her early 70’s woman with KL2 (two images on the right). *Abbreviations: Corr.EI, corrected echo intensity; FB, femoral bone; KL grade, Kellgren Lawrence grade; MTFJ, medial tibiofemoral joint; RF, rectus femoris; SCF, subcutaneous fat; VM, vastus medialis; WORMS, whole-organ magnetic resonance imaging score*.

**Figure 4.**
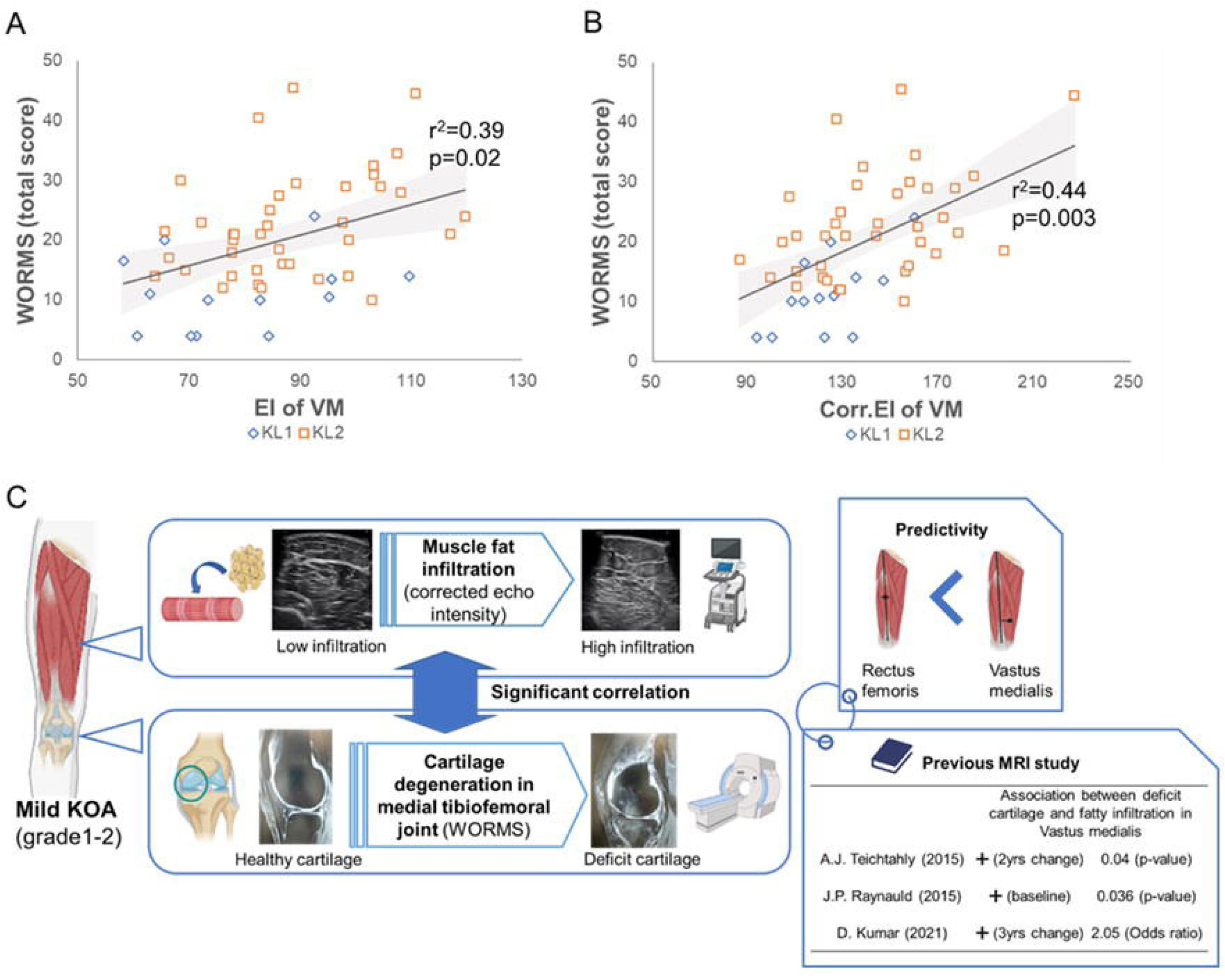
Relationship between quadriceps muscle quality and structural abnormalities. Scatter plot and regression line between WORMS total score and EI of VM (**A**) and corrected EI of VM (**B**). The regression line was calculated from total data (KL1 and KL2). This study showed that individuals with poor muscle quality in the VM, rather than RF, displayed compromised cartilage integrity in the medial tibiofemoral joint at an earlier stage of the disease (**C**). These results of present ultrasound study were consistent to previous MRI studies that assessed correlation between increased intramuscular fat and cartilage loss (**C**). *Abbreviations: Corr.EI, corrected echo intensity; EI, echo intensity; KL, Kellgren and Lawrence; MRI, magnetic resonance imaging; RF, rectus femoris; VM, vastus medialis; WORMS, whole-organ magnetic resonance imaging score; yrs, years*.

**Table 2.** Association between WORMS total score and VM and RF muscle quality.

| VM (corr.EI) vs WORMS (total score) |  |  |  |  |  |
| --- | --- | --- | --- | --- | --- |
| Variables | Coefficient | 95% CI | Df | t value | p value |
| (Intercept) | -17.477 | -37.552, 2.644 | 45.807 | -1.660 | 0.104 |
| Corr. EI (VM) | 0.146 | 0.06, 0.231 | 46.523 | 3.269 | 0.002 |
| Muscle thickness (VM) | -2.45 | -7.118, 2.225 | 45.199 | -1.008 | 0.319 |
| Age | 0.34 | 0.096, 0.583 | 43.782 | 2.644 | 0.011 |
| Sex | -1.074 | -6.288, 4.127 | 45.466 | -0.392 | 0.697 |
| RF (corr.EI) vs WORMS (total score) |  |  |  |  |  |
| (Intercept) | -24.942 | -52.471, 2.389 | 44.402 | -1.744 | 0.088 |
| Corr.EI (RF) | 0.054 | -0.048, 0.157 | 46.862 | 1.014 | 0.316 |
| Muscle thickness (RF) | 5.712 | -2.687, 14.151 | 46.07 | 1.386 | 0.172 |
| Age | 0.473 | 0.205, 0.742 | 44.046 | 3.362 | 0.002 |
| Sex | -5.727 | -11.54, 0.062 | 46.113 | -1.904 | 0.063 |

Regression coefficients and 95% CIs of corrected EI (continuous variable) were calculated, demonstrating that the corrected EI of VM is more predictive than corrected EI of RF for the WORMS total score (continuous variable) while simultaneously including (1-step model), muscle thickness (continuous variable), age (continuous variable) and sex.
*Abbreviations: 95% CI, 95% confidence interval; Corr. EI, corrected echo intensity; Df,* *degrees of freedom; EI, echo intensity; RF, rectus femoris; VM, vastus medialis; WORMS,* *whole-organ magnetic resonance imaging score.*

Because joint structural abnormalities were most frequently identified in the articular cartilage in the medial tibiofemoral joint (**Figure 2**), we performed a sensitivity analysis focusing on the WORMS cartilage score in the medial tibiofemoral joint as an outcome variable. The results revealed that poor muscle quality of the VM was significantly associated with more cartilage abnormalities in the medial tibiofemoral joint (**Table S1**). When we added the lower limb alignment (i.e., corrected FTA) or BMI as a covariate to the generalized linear mixed model, poor muscle quality of VM still displayed a significant association with cartilage in medial tibiofemoral joint (**Table S2, S3**). Collectively, these findings indicate that individuals with poor VM muscle quality demonstrate disrupted cartilage integrity in the medial compartment of the knee, regardless of the degree of static knee alignment or obesity. Our results identified by ultrasound reinforce those previously identified by MRI demonstrating that increased cartilage damage is associated with increased fat infiltration in the quadriceps, particularly VM muscle(9, 34–36) (**Figure 4C**).

## DISCUSSION

Establishment of ultrasound-based prognostic disease marker at the early stages of KOA represents a critical need in the field to better manage the progression of OA. As a first step to address this need, this study tested the hypothesis that poor quadriceps muscle quality, as assessed by ultrasound, is associated with greater structural abnormalities in participants with early stages of KOA. We found that individuals with poor VM muscle quality demonstrated significantly greater structural abnormalities in the knee, even after adjustment for muscle thickness, age, and sex. A subsequent sensitivity analysis identified the same relationship when we focused on structural abnormalities in the articular cartilage at the medial tibiofemoral joint, a finding consistent with medial KOA(33). Notably, these relationships were not significant in the RF muscle, suggesting that the identified relationship between poor quadriceps muscle quality and structural abnormalities occurs in a muscle-specific manner. Importantly, in these analyses, EI corrected for subcutaneous fat improved the predictive ability for structural integrity. Taken together, poor muscle quality in the VM assessed by ultrasound may be a sensitive marker for disrupted cartilage integrity and development of KOA.

Analytical ultrasound imaging is increasingly recognized as a cost-efficient and reliable tool to assess fat content in skeletal muscle with many benefits over the two gold standard methods, MRI and CT. Ultrasound-based assessment is also attractive over other measures of muscle dysfunction, such as manual muscle strength testing, given it is less influenced by pain evoked by the measurement procedure (37, 38). Here, we evaluated muscle quality by ultrasound using a previously validated scale, EI. We found that quadriceps EI was an effective predictor of individuals with compromised cartilage integrity. Our results are in line with longitudinal studies demonstrating that VM fat content as evaluated by MRI predicted greater cartilage loss and bone marrow lesions(9, 35, 36). The similarities between the different devices supports the use of ultrasound as a promising tool to predict disrupted cartilage integrity. The utility of ultrasound-based assessment is further enhanced by its capacity to identify individuals at an earlier stage of KOA when compared to conventional radiography, which does not adequately capture the onset or progression of cartilage damage.

Early KOA represents a critical window of opportunity to restore joint integrity before the onset of debilitating pain and functional limitations, which often require invasive interventions(2). The findings of this study suggest that implementing ultrasound in clinical practice may help to identify people with minimal cartilage damage who are at risk of developing more severe KOA.

Through ultrasound assessments of individual quadriceps muscles, this study also found that the VM displayed a greater predictive ability for compromised cartilage integrity than RF. This was also consistent with previous longitudinal studies using MRI assessments, which revealed the significant relationship between cartilage loss and increased fat infiltration in the VM, but not other quadriceps muscles (i.e., RF, vastus lateralis or vastus intermedius muscles)(36). While the underlying role of the VM in cartilage damage is unclear, these observations may reflect the unique biomechanical role of the VM muscle. The VM primarily acts to stabilize the patellofemoral joint by maintaining medial tracking of the patella during loading(39, 40). The VM therefore contributes to joint stability and load distribution of the knee in the medial-lateral plane with the potential to affect the integrity of articular cartilage at medial tibiofemoral joint(41). Another possible explanation for muscle-specific association with cartilage loss is metabolic link between adipose tissue in VM and cartilage homeostasis. Fat is an endocrine organ that releases inflammatory cytokines such as tumor necrosis factor alpha and interleukin-6(42), which have been implicated in the pathophysiology of KOA(43). The role of adipose-cartilage molecular crosstalk was clearly presented by the recent animal study demonstrating that fat deficiency protected knee joints from spontaneous or post-traumatic murine KOA(44). While no study has addressed the differential role of fat in the VM versus RF muscles, the close anatomic proximity of VM and medial tibiofemoral joint might explain, at least in part, the observed relationship between fat infiltration in VM and cartilage integrity. The current study further supports the need for studies investigating the crosstalk between adipose in quadriceps muscle, particularly in the VM, and cartilage.

Although this study provides a new perspective on a sensitive marker for early stage of KOA, it has limitations. First, this study did not consider physical activity levels as a potential confounder(45, 46). Reduced physical activity in daily life, commonly seen in people with KOA, increases accumulation of inter and intramuscular adipose tissue(47). Also, the cross-sectional nature of the study design limited determination of the causal relationship between poor VM muscle quality and compromised structural integrity.

Nevertheless, this study has several strengths. First, we applied subcutaneous fat correction to ultrasound-based muscle quality assessment, as recommended in previous reports(19, 20). This method was used to adjust for ultrasound wave attenuation due to subcutaneous fat(48). The corrected EI improved the predictive ability of ultrasound-based signal intensity for structural integrity, and, therefore, this metric should be considered for use in clinical practice over EI. Second, this study used MRI to assess multiple tissues involved in OA pathology. This allowed for the assessment of the relationship between quadriceps muscle quality and structural integrity as stratified according to tissue type, such as cartilage, subchondral bone, and synovium. All of these are considered to influence each other via molecular crosstalk, which contributes to the pathophysiology of early KOA(49). Finally, this study analyzed patients with symptomatic mild KOA who are generally considered to be at high risk of disease progression. Targeting this population provides an enhanced identification of patients who might benefit from treatment for KOA to prevent disease progression.

Overall, this study addressed an existing knowledge gap by demonstrating that poor muscle quality in the VM muscle in early-stage KOA is related to structural abnormalities. Further, findings of the current study support the use of ultrasound as a reliable tool for assessing VM muscle quality as a sensitive marker of earlier stage of KOA. As such, our study has the potential to represent an important first step in the development of future longitudinal studies designed to clarify the role of muscle quality, and, ultimately, refine clinical practice to target disease progression at an earlier stage.

## Contributors

(1) AH and HI were the main contributors to the conception and design of the study; AH, TI, and YM were the main contributors to the acquisition of data; AH, ED, FA, and HI contributed to the analysis and interpretation of data.
(2) AH, ED, FA, and HI drafted the manuscript, and AH, ED, FA, and HI critically revised the manuscript.
(3) All authors have provided final approval for the version to be submitted.

## Competing interests

The authors have no conflicts of interest to declare.

## Data Availability

All data produced in the present study are available upon reasonable request to the authors.

## Acknowledgments

We thank the members of the Physical Therapy and Radiology Department of Matsuda Orthopedic Clinic for extending support in data collection.

## Notes

### Competing Interest Statement

The authors have declared no competing interest.

### Funding Statement

This study did not receive any funding.

### Author Declarations

Ethics committee/IRB of Medical Corporation Nagomi gave ethical approval for this work.

### Summary of Updates

Section on introduction, result and discussion updated to clarify the muscle-specific differences.

