## Supplementary material for "Fat infiltration in the vastus medialis implicates joint structural abnormalities in early-stage symptomatic knee osteoarthritis": TableS1-S3

**The PDF file includes:**

Table S1. Sensitivity analysis for the assessment of the muscle-specific differences in cartilage integrity and muscle quality

Table S2. Sensitivity analysis for the assessment of the correlation between cartilage integrity and muscle quality with adjusted by static alignment

Table S3. Sensitivity analysis for the assessment of the correlation between cartilage integrity and muscle quality with adjusted by body mass index

This supplementary material has been provided by the authors to give readers additional information about their work

Table S1. Sensitivity analysis for the assessment of the muscle-specific differences in cartilage integrity and muscle quality.

| VM (corr.EI) vs WORMS (cartilage score in the medial tibiofemoral joint) | | | | | |
| --- | --- | --- | --- | --- | --- |
| Variables | Coefficient | 95% CI | Df | t value | p value |
| (Intercept) | -6.127 | -14.402, 2.16 | 45.832 | -1.407 | 0.166 |
| Corr. EI (VM) | 0.066 | 0.031, 0.102 | 46.932 | 3.57 | 0.0008 |
| MT (VM) | 0.832 | -1.09, 2.753 | 45.491 | 0.828 | 0.412 |
| Age | 0.028 | -0.072, 0.129 | 44.279 | 0.538 | 0.593 |
| Sex | -0.003 | -2.166, 2.151 | 45.538 | -0.003 | 0.998 |
| RF (corr.EI) vs WORMS (cartilage score in the medial tibiofemoral joint) | | | | | |
| (Intercept) | -2.303 | -13.584, 8.98 | 45.037 | -0.387 | 0.70 |
| Corr. EI (RF) | 0.032 | -0.011, 0.075 | 46.324 | 1.420 | 0.162 |
| MT (RF) | 0.484 | -2.923, 3.896 | 46.996 | 0.277 | 0.783 |
| Age | 0.06 | -0.05, 0.171 | 44.05 | 1.036 | 0.306 |
| Sex | -1.253 | -3.634, 1.128 | 45.061 | -0.998 | 0.324 |

Regression coefficients and 95% CIs of corrected EI (continuous variable) were calculated to indicate their predictive ability for the total WORMS cartilage score (continuous variable) or WORMS cartilage score in the medial tibiofemoral joint (continuous variable) while simultaneously including (1-step model) vastus medialis MT (continuous variable), age (continuous variable) and sex.

*Abbreviations: 95% CI, 95% confidence interval; Corr.EI, corrected echo intensity; Df, degrees of freedom; EI, echo intensity; MT, muscle thickness; RF, rectus femoris; VM, vastus medialis; WORMS, whole-organ magnetic resonance imaging score.*

Table S2. Sensitivity analysis for the assessment of the correlation between cartilage integrity and muscle quality with adjusted by static alignment.

| VM (corr.EI) vs WORMS (cartilage score in the medial tibiofemoral joint) | | | | | |
| --- | --- | --- | --- | --- | --- |
| Variables | Estimate | 95% CI | Df | t value | p value |
| (Intercept) | 10.069 | -74.993, 95.351 | 44.883 | 0.222 | 0.825 |
| Corr. EI (VM) | 0.067 | 0.031, 0.102 | 45.931 | 3.551 | 0.0009 |
| MT (VM) | 0.922 | -1.054, 2.891 | 44.564 | 0.882 | 0.383 |
| Age | 0.025 | -0.076, 0.127 | 43.533 | 0.471 | 0.64 |
| Sex | 0.317 | -2.412, 3.037 | 45.09 | 0.218 | 0.828 |
| Corr. FTA | -0.089 | -0.558, 0.378 | 44.932 | -0.359 | 0.721 |

WORMS cartilage score in the medial tibiofemoral joint (continuous variable) while simultaneously including (1-step model) age (continuous variable), vastus medialis MT (continuous variable), sex and corrected FTA (continuous variable).

*Abbreviations: 95% CI, 95% confidence interval; Corr. FTA, corrected femur-tibia angle; Corr. EI, corrected echo intensity; Df, degrees of freedom; MT, muscle thickness; VM, vastus medialis; WORMS, whole-organ magnetic resonance imaging score.*

Table S3. Sensitivity analysis for the assessment of the correlation between cartilage integrity and muscle quality with adjusted by body mass index.

| VM (corr.EI) vs WORMS (cartilage score in the medial tibiofemoral joint) | | | | | |
| --- | --- | --- | --- | --- | --- |
| Variables | Estimate | 95% CI | Df | t value | p value |
| (Intercept) | -6.788 | -15.869, 2.299 | 43.826 | -1.402 | 0.168 |
| Corr. EI | 0.062 | 0.02, 0.104 | 44.925 | 2.799 | 0.008 |
| MT (VM) | 0.767 | -1.189, 2.72 | 44.702 | 0.744 | 0.461 |
| Age | 0.031 | -0.07, 0.133 | 43.679 | 0.576 | 0.568 |
| Sex | -0.111 | -2.369, 2.125 | 45.314 | -0.094 | 0.926 |
| BMI | 0.048 | -0.227, 0.326 | 45.364 | 0.329 | 0.744 |

WORMS cartilage score in the medial tibiofemoral joint (continuous variable) while simultaneously including (1-step model) age (continuous variable), vastus medialis MT (continuous variable), sex and BMI (continuous variable).

*Abbreviations: 95% CI, 95% confidence interval; BMI, body mass index; Corr. EI, corrected echo intensity; Df, degrees of freedom; MT, muscle thickness; VM, vastus medialis; WORMS, whole-organ magnetic resonance imaging score.*
